# Altered Neural Oscillations Underlying Visuospatial Processing in Cerebral Visual Impairment (CVI)

**DOI:** 10.1101/2022.10.10.22280895

**Authors:** Alessandra Federici, Christopher R. Bennett, Corinna M. Bauer, Claire E. Manley, Emiliano Ricciardi, Davide Bottari, Lotfi B. Merabet

**Affiliations:** IMT School for Advanced Studies. Lucca, 55100 Lucca, Italy; The Laboratory for Visual Neuroplasticity, Department of Ophthalmology, Massachusetts Eye and Ear, Harvard Medical School, Boston, MA, 02114, USA

**Keywords:** Neural Oscillations, Cerebral Visual Impairment, Feedforward, Feedback, Visual Search, Visuospatial

## Abstract

Visuospatial processing deficits are commonly observed in individuals with cerebral visual impairment (CVI) and even in cases where visual acuity and visual field functions are intact. CVI is a brain-based visual disorder associated with the maldevelopment of central visual pathways and structures. However, the neurophysiological basis underlying higher-order perceptual impairments in this condition has not been clearly identified, which in turn posits limits on rehabilitative interventions. Using combined eye tracking and electroencephalography (EEG) recordings, we assessed the profile and performance of visual search on a naturalistic virtual reality (VR)-based task. Participants with CVI and controls with neurotypical development were instructed to search, locate, and fixate a specific target placed among surrounding distractors at two levels of task difficulty. Analyzing evoked (time or phase-locked) and induced (not time or phase-locked) components of EEG activity allowed for feedforward and feedback processing mechanisms to be uncovered. We found that visual search performance in CVI was impaired compared to age-matched controls with neurotypical development (as indexed by outcomes of success rate, reaction time, and gaze error). Analysis of neural oscillations across a broad frequency band [4-55 Hz] revealed markedly reduced early-onset evoked theta [4-6 Hz] activity (within 0.5 sec) regardless of task difficulty. Moreover, while induced alpha activity increased with task difficulty in controls, this modulation was absent in the CVI group providing a potential neural correlate for their deficits with visual search and distractor suppression. Finally, CVI participants also showed an overall delayed and sustained induced gamma response [30-45 Hz]. We conclude that impaired visual search performance in CVI is associated with substantial alterations across a wide range of neural oscillation frequencies. This includes both evoked and induced components related to feedforward and feedback processing and implicating local and distributed levels of neural processing.

## Introduction

Cerebral visual impairment (CVI) is a brain-based visual disorder associated with damage or maldevelopment of retrochiasmatic visual processing areas ^1,2^, and is the leading individual cause of pediatric visual impairment in developed countries ^3-5^. Early neurological damage (e.g., hypoxic-ischemic injury, trauma, infection, and genetic/metabolic disorders; ^6^) to visual pathways and higher-order processing areas of the brain are commonly very heterogeneous and thus contribute to the broad clinical presentation of visual impairments reported in this population ^7-10^. Reduced visual acuity, visual field function, contrast sensitivity, as well as impaired ocular motor functions are all commonly observed in CVI. However, visual perceptual deficits related to higher-order visuospatial and visual attention processing are also highly prevalent ^4,7,8,11^, even in cases where visual acuity and visual field functions are at normal or near-normal levels ^12,13^. Interestingly, individuals with CVI often report difficulties interacting with complex and cluttered visual scenes ^9,14-16^. For example, an individual may easily identify a favorite toy or recognize a familiar person when presented in isolation yet have difficulties finding them in a cluttered toybox or in a crowd ^9,10,15^. Taken together, these associated visual deficits can have a substantial impact upon an individual’s functioning, independence, and well-being ^17^. Despite representing a significant public health concern, the neurophysiological basis of CVI remains poorly understood. One important and unresolved question is: are visuospatial processing deficits mainly driven by aberrant early stage (i.e., feedforward) sensory input, or are they the result of impaired higher-order (i.e., feedback) visual processing mechanisms? Answering this question could be useful in identifying potential biomarkers of CVI, and potentially help inform the design of specific training strategies.

In clinical practice, visual-evoked potential (VEP) recordings have been used to assess the integrity of afferent visual pathways and functions (such as visual acuity), particularly in the case of infants and/or nonverbal patients who may not be able to undergo formal ophthalmic testing ^3,18,19^; see ^20^ for review). VEPs are typically measured by averaging neural activity in response to passive viewing of simple visual stimuli such as flashes of light or patterns (e.g., gratings and checkerboards). However, clinical VEPs remain limited with respect to characterizing processing within visual areas beyond primary visual cortex (V1) and the underlying neurophysiological basis of higher-order visual processing deficits observed in CVI. That is, the recording of typical VEP responses do not rule out the presence of higher-order processing deficits. In this regard, the analysis of neural oscillatory activity may be more informative with respect to how information is encoded, transferred, and integrated within the brain ^21,22^. Neural oscillations are considered to play a crucial role in perception by integrating signals across multiple brain regions ^22-25^. By analyzing a neural network’s intrinsic ability to resonate and oscillate at multiple frequencies, neural activity associated with both sensory driven (i.e., evoked; time or phase locked to stimulus onset) as well as processes related to the integration of sensory inputs (i.e., induced; not time or phase locked to stimulus onset) can be characterized ^26-28^. Furthermore, distinct components of neural oscillations are known to be associated with different types of processing with respect to the direction of information flow ^25,29^. Alterations in evoked activity likely reveal impairments with feedforward information processing along the thalamocortical pathway, while abnormal induced activity is consistent with deficits in feedback processing associated with cortico-cortical connectivity (see ^25,30^). This approach has been used to identify potentially important biomarkers of aberrant oscillatory activity in a variety of neurodevelopmental conditions including autism spectrum disorder ^31^, dyslexia ^32^ and other language-learning impairments ^33-35^.

In a preliminary study, we recorded electroencephalography (EEG) signals while CVI participants carried out a virtual reality (VR)-based visual search task ^36^. Using visual search as a proxy for visuospatial and visual attention processing abilities ^37^, we found that impaired search performance in CVI was associated with a dramatic reduction in alpha desynchronization activity ^36^. Given that alpha desynchronization is an important neural signal associated with visuospatial attention and distractor suppression ^38^, these findings provided early evidence of altered neural processing in relation to higher-order visual perceptual deficits in this population. In the present study, we aimed to extend these findings by further investigating neural oscillatory activity in relation to manipulating visual search task difficulty as defined by the presence of clutter in a visual scene (i.e., complexity). By investigating a broad frequency range [4-55 Hz] of neural oscillations, as well as their evoked and induced related activity associated with overt visual search performance, we could better characterize the contribution of early stage (i.e., feedforward) and higher-order (i.e., feedback) visual processing mechanisms in CVI. As a secondary aim, we also investigated putative associations between EEG activity and the clinical profile such as visual acuity as well as available structural morphometry data (MRI) in our CVI participants.

## Materials and methods

### Participants

A total of twenty-six individuals were recruited in this study. Ten participants were previously diagnosed with CVI (three females, mean age: 17.5 years old ± 2.59 SD) by experienced clinicians specializing in neuro-ophthalmic pediatric care (see supplementary information details regarding diagnosis criteria). All participants with CVI had visual impairments related to pre- or perinatal neurological injury and/or neurodevelopmental disorders. Causes of CVI included hypoxic-ischemic injury related to prematurity (including periventricular leukomalacia; PVL), hypoxic/ischemic encephalopathy (HIE), seizure disorder, as well as genetic and metabolic disorders. Four out of the 10 CVI participants were born preterm (i.e., prior to 37 weeks gestation). Associated neurodevelopmental comorbidities included spastic and dystonic cerebral palsy. Best corrected visual acuities in the better seeing eye ranged from 20/20 to 20/60 Snellen (0.0 to 0.5 logMAR equivalent). CVI participants were also categorized according to previous functional criteria ^1^ and the distribution of our study population was limited to categories 2 and 3 (definition of category 2: 40% defined as “have functionally useful vision and cognitive challenges”, and category 3: 60% defined as “functionally useful vision and who can work at or near expected academic level for their age group”). For a full list of CVI participant details, see Table 1. All CVI participants had a level of visual acuity, intact visual field function within the area corresponding to the visual stimulus presentation, as well as fixation and binocular ocular motor function sufficient for the purposes of completing the behavioral task requirements (including eye tracking calibration; see supplementary information for further details). Exclusion criteria included any evidence of oculomotor apraxia, intraocular pathology (other than mild optic atrophy), uncorrected strabismus, a visual field deficit corresponding to the area of testing, uncontrolled seizure activity, as well as cognitive deficits precluding the participant from understanding the requirements of the study.

Structural MRIs from CVI participants were assessed for brain lesion severity according to a reliable and validated semi-quantitative scale. The scoring procedure has been described in detail elsewhere ^39^. Briefly, raw scores for each brain structure (e.g., lobe, subcortical structures, corpus callosum, and cerebellum) were calculated and summed to provide a global (total of cortical and subcortical) lesion score, whereby a higher score is indicative of a greater degree of brain injury (representative images from CVI participants are shown in supplementary figure S1).

Sixteen individuals with neurotypical development (five females, mean age: 19.2 years old ± 2.14 SD) were recruited as comparative controls. Control participants had normal or corrected to normal visual acuity and no previous history of any ophthalmic (e.g., strabismus, amblyopia) or neurodevelopmental (e.g., epilepsy, attention deficit disorder) conditions. The control and CVI groups were not statistically different with respect to mean age [t(24) = 1.806, p = 0.084].

Formal written consent was obtained from all the participants and a parent/legal guardian (in the case of a minor). The study was approved by the investigative review board of the Massachusetts Eye and Ear, Boston MA, USA and carried out in accordance with the Code of Ethics of the World Medical Association (Declaration of Helsinki) for experiments involving humans.

### Visual Search Task Design and Eye Tracking

The behavioral task was a static object visual search based on a previously designed desktop VR-based naturalistic environment called the “virtual toybox” (complete details regarding task design can be found in ^40^, see also ^41^). Briefly, the task represented a simulated rendering of a toybox viewed from an overhead, first-person perspective. Participants were instructed to search, locate, and fixate a specific target toy (a blue truck) placed randomly among surrounding toys (and without overlap) serving as distractors in a 5 × 5 array. The target toy remained constant and was unique with respect to shape and color so as to create a “pop out” effect (akin to feature search). The arrays of toys were presented in a trial-by-trial fashion and pseudorandom order. Two levels of task difficulty (“low” and “high”) were used. In the low task condition, the target toy was surrounded by one unique distractor toy presented on a uniform background. In the high task condition, the target was placed alongside nine unique distractor toys and superimposed upon a cluttered background scene designed to make target recognition more difficult (see figure 1). At a viewing distance of 60 cm, the virtual toybox subtended approximately 38 × 38 deg of visual angle. An equal number of low and high task conditions were presented. A trial consisted of viewing the toybox scene for 2 sec followed by 1 sec of a blank gray screen with a central fixation target. This was repeated 50 times per run, and 4 runs were collected (total of 200 trials). Each run lasted less than 2.5 min, with a brief rest period between each run.

**Figure 1.**
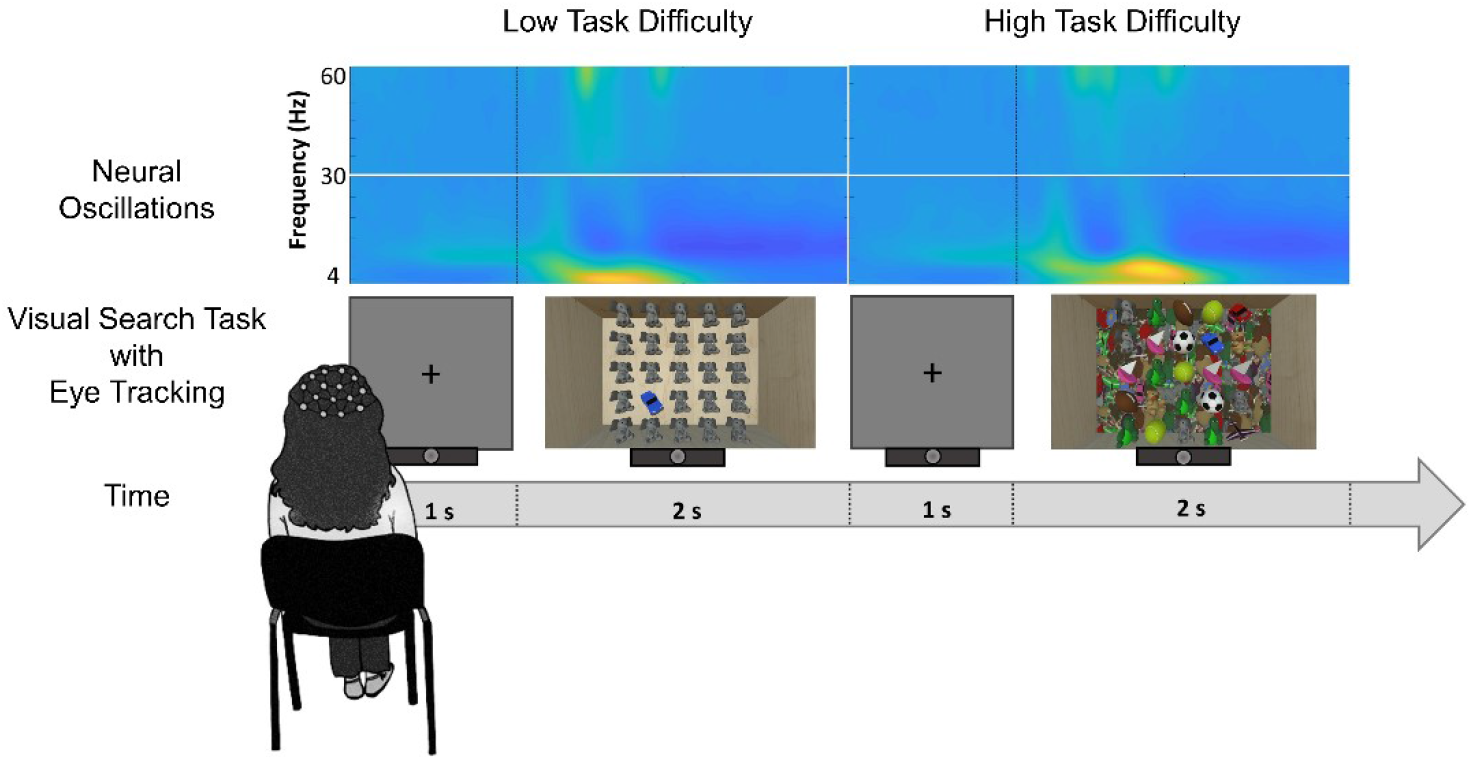
Experimental Design. Illustrative figure showing a participant performing the visual search task while eye gaze patterns and EEG related neural activity are recorded (putative neural oscillatory activity is shown in the top row). Two trials are shown, one for each visual task condition (low and high task difficulty). Between groups statistical significance levels: ***p<0.001.

### Visual Search Performance Outcomes and Behavioral Data Analysis

Participants were seated comfortably in a quiet room, 60 cm in front of a 27” LED monitor (ViewSonic 27” Widescreen 1080p; 1920 × 1080 resolution). Visual search performance was quantified using three objective outcomes based on captured eye tracking data. Success rate measured the percentage of trials in which the participant found and fixated the target in a given run. Successful fixation was defined as sustained gaze that remained within the outer contour of the target for a minimum time of 400 msec. Reaction time corresponded to the time needed to find the target on the screen and was defined as the first moment the participant’s gaze arrived within the outer contour of the target and remained fixated for at least 400 msec ^40,41^. Gaze error was a continuous measure of a participant’s ability to locate and accurately fixate the target ^40,41^. This accuracy measure was defined as the distance between the center of the target and the participant’s gaze position and computed based on the sampling rate of the eye-tracker (90 Hz).

Statistical analyses of behavioral data were carried out using a mixed-model repeated measure ANOVA (with group as a between factor and condition as a within factor) and conducted separately for each of the visual search outcome measures. All statistical analyses were performed using IBM SPSS v 26 Statistics Software Package, SAS Studio, and MATLAB Statistics Toolbox.

### Electrophysiological Data Acquisition, Signal Processing, and Analysis

Electrophysiological data were acquired while participants performed the visual search task using a wireless 20-channel Enobio system (Neuroelectrics, Barcelona, Spain; sampling rate of 500 Hz). This system was chosen for its simplified montage and setup procedure to minimize participant discomfort. After the recording cap was placed on the participant, conductive gel was added to each electrode site in order to enhance conductivity between the electrode and scalp surface. Once the montage was ready, signal quality was verified at each channel and additional conductive gel was added if necessary to ensure adequate signal recording quality. Recording electrodes were made of a solid gel material and each channel fed to a wireless (Bluetooth®) transmitter. Electrodes were arranged according to the standard 10-20 international system and covered the entire cap. An additional reference channel was connected using a clip placed on the participant’s right ear lobe. Signals were captured on-line and recorded by acquisition software running on a desktop computer located within 1 m of the participant.

EEG signal preprocessing was performed by implementing a validated pipeline to clean the data and promote reproducibility ^42,43^. Offline, each participant’s continuous dataset was low-pass (cut-off at 40 Hz, filter order 50) and high-pass (cut-off at 1 Hz, filter order 500) filtered using a Hanning filter ^44^, down sampled to 250 Hz, segmented into consecutive 1 sec epochs, and cleaned by using a joint probability algorithm (epochs displaying activity with a joint probability across all channels exceeding a 3 SD threshold were removed; ^45^). An independent component analysis (ICA) based on the extended Infomax algorithm ^46-48^ was then performed, and the resulting weights were applied to the raw continuous unfiltered data ^42,43^. Components associated with stereotypical eye blinks were identified and removed using a semiautomatic procedure (CORRMAP, ^49^). The template for blink components was chosen from the control group to ensure the selection of a typical artifact, and this template was used to find blink components across all participants (mean components removed for each individual in the CVI group = 0.9 ± 0.32 SD and control group = 0.94 ± 0.25 SD; see supplementary figure S2 for the topography of each component removed in each participant). All EEG activity associated with eye movements was retained due to the overt nature of the behavioral task and to avoid the risk of removing associated brain activity. Datasets were then low-pass (60 Hz, filter order 26) and high-pass (0.1 Hz, filter order 1000) filtered with a Hanning filter. Each continuous EEG signal was then segmented into epochs of 3 sec (from −1 to 2 sec with respect to the onset of the visual stimulus) and split between the low and high levels of task difficulty. Noisy channels were identified based on visual inspection, and then interpolated using a spherical spline (mean interpolated electrodes per subject in the CVI group: 1.7 ± 1.06 SD and in the control group: 1.1 ± 0.84 SD). Following visual inspection, we rejected epochs exceeding a threshold of 150 µV within the [-0.5 - 1 sec] time window representing contaminated portions of the EEG signal. Remaining epochs were further cleaned using a joint probability across channels with a threshold of 3 SD ^45^. In total, the mean epochs rejected per subject in the CVI group were 12.7% ± 6.6 SD for low and 11.2% ± 5.6 SD for the high task difficulty trials and in the control group these were 8.4% ± 2.9 SD for low and 8.6% ± 2.4 SD for the high task difficulty trials. Note that no statistically significant difference between groups emerged (all p values > 0.07). Finally, we re-referenced the data to the average and a 40 Hz low-pass filter (order 50) was applied only before the computation of the event-related potential (ERP). All these steps were performed with EEGLAB software ^50^. Data were then imported in Fieldtrip ^51^ to perform time-frequency decomposition, ERP, and statistical analyses. We also extracted ERPs separately for the low and high levels of task difficulty for each group. Data were baseline corrected, using a time window between −0.1 to 0 sec before the visual stimulus presentation as baseline.

Primary analysis of acquired EEG activity was focused on oscillatory activity associated with visual search behavior. To this end, time-frequency decomposition of the EEG data was performed separately for the low and high levels of task difficulty within each group (CVI and controls). At the single participant level, we first extracted the induced power at each single trial after subtracting from each epoch the non-filtered ERP (i.e., computed for each participant without 40 Hz low-pass filtering) to remove the evoked activity from each trial. Single-trial time-frequency decomposition was computed at each electrode and separately for low [2-30 Hz] and high [30-55 Hz] frequency ranges. The higher frequency range was investigated up to 55 Hz to avoid line noise contamination (i.e., 60 Hz). Within the lower frequency range [2-30 Hz], oscillatory power was estimated using a Hanning taper with a frequency-dependent window length (4 cycles per time window) in steps of 2 Hz. Within the higher frequency range [30-55 Hz], oscillatory power was estimated using a Multitapers method with a Slepian sequence as tapers (in steps of 5 Hz) with a fixed-length time window of 0.2 sec and fixed spectral smoothing of ± 10 Hz. For both low and high frequency ranges, power was extracted over the entire epoch (from −1 to 2 sec) and in steps of 0.02 sec. Then, the average across trials was computed at the single-subject level for both conditions (i.e., low and high levels of task difficulty) and for the lower and higher frequency range. The resulting oscillatory activity was baseline-corrected to obtain the relative change with respect to the baseline period. For the low frequency range, the baseline was set between −0.7 and −0.3 sec, while for the high frequency range, this was set between −0.2 and −0.1 sec. Lower frequencies (i.e., having a longer cycle) required a wider baseline window for appropriate estimation of slow oscillations and farther away from 0 to avoid temporal leakage of post-stimulus activity into the pre-stimulus baseline. The same procedure was implemented without ERP subtraction from single trials to estimate the total power from which the baseline-corrected evoked power was computed by subtracting the baseline-corrected induced power.

The same statistical approach was applied for all extracted electrophysiological measures. To investigate whether there was an interaction effect between the factors of group and task difficulty on oscillatory activities (i.e., evoked and induced components; low and high frequency ranges) and ERPs, we estimated the neural response change between conditions (differential activity computed from high minus low levels of task difficulty) within each group. Non-parametric cluster-based permutation (without a priori assumptions) was performed between the CVI and control groups based on the difference between conditions (high-low). The Monte Carlo method with 1000 random permutations was applied and cluster-level statistics were calculated taking the sum of the t-values within every cluster (minimum neighbor channel = 1; alpha level of 0.05, two-tailed to account for positive and negative clusters). Thus, identified clusters were considered significant at p < 0.025 (alpha=0.05, two-tailed). For ERPs, the cluster-based permutation analyses were performed across all electrodes and time points within the time window [0-0.5 sec]. For time-frequency decomposition, statistical analyses were performed across all electrodes for time points within the time window [0-1] sec and across all frequencies within low [4-30 Hz] and high [30-55 Hz] ranges, and separately performed for induced and evoked oscillatory activities.

If a significant interaction effect emerged, we assessed the difference between conditions within each group (i.e., low vs. high task conditions in controls and low vs. high conditions in the CVI group), and the difference between the two groups within each condition (i.e., controls vs. CVI in the high task condition and controls vs. CVI in the low task condition). These effects were all investigated with the same cluster-based permutation analysis approach and parameters described above (p<0.025, alpha=0.05, two-tailed). In the absence of an interaction effect, we collapsed the two conditions, performing the average between the low and high task difficulty conditions. The same cluster-based permutation analysis approach was performed on neural activity in the control and CVI groups (p<0.025, alpha=0.05, two-tailed).

### Volumetric analysis of thalamic nuclei and pericalcarine cortex

Structural morphometry data were available from a subset of participants with CVI (n=9). Of these, 6 were scanned using a T1-weighted anatomical scan (TE 2.9 msec, TR 6.5 msec, flip angle 8°, isotropic 1 mm voxel size) acquired with a 32-channel phased array head coil (Philips 3T Elition X scanner). An additional 3 participants with CVI were scanned as part of a previous protocol (TE 3.1 msec, TR 6.8 msec, flip angle 9°, isotropic 1 mm voxel size) using an 8 channel phased array head coil (Philips 3T Intera Achieva). The volumes of predetermined thalamic nuclei (i.e., the lateral geniculate nucleus (LGN) and pulvinar) and the pericalcarine cortex were quantified for each subject in anatomical space using the FreeSurfer 7.2.0 (https://surfer.nmr.mgh.harvard.edu) ^52-55^. Segmentations for subcortical and cortical ROIs were derived from a probabilistic thalamic atlas ^56^ and the Desikan-Killiany atlas ^57^, respectively.

### Correlation Analyses

We investigated putative associations between EEG activity and the clinical profile (i.e., age, visual acuity, and global lesion classification) as well as available structural morphometry data (MRI) of our CVI participants using Spearman rank correlations followed by correction for multiple comparisons using False Discovery Rate (FDR) ^58^. Correlations between EEG activity and the volumes of the LGN, pulvinar, and primary visual cortex were evaluated following adjustment for estimated total intracranial volume using residuals.

### Data availability

The data that support the findings of this study are available from the corresponding author, upon reasonable request and pending investigative review board approval.

## Results

### Visual Search Task Performance

We examined visual search performance on the effect of varying task difficulty with respect to all outcomes of interest. We conducted a series of mixed ANOVAs with group (CVI and control) as between-subject factor and task difficulty (high and low) as within-subject factor. In general, for all outcomes of interest (including success rate, reaction time, and gaze error), we found a significant group difference between controls and CVI, with the CVI group having an overall impairment in visual search performance. A task difficulty difference between low and high conditions was found for reaction time and gaze error. No interaction effect between the task difficulty and group conditions emerged (see below).

CVI participants showed a lower mean success rate compared to controls. A mixed ANOVA revealed that the CVI group had a significantly lower success rate (low: 84.06 % ± 21.18 SD, high: 81.30 % ± 22.24 SD) than controls (low: 99.22 % ± 2.15, high: 98.28 % ± 3.84) [F(1,24) = 9.141 p = 0.006, ηp2 = 0.276]. There was no effect of difficulty level [F(1,24) = 2.36, p = 0.137, ηp2 = 0.090] and no interaction effect [F(1,24) = 0.57, p = 0.458, ηp2 = 0.023] (figure 2 A).

**Figure 2.**
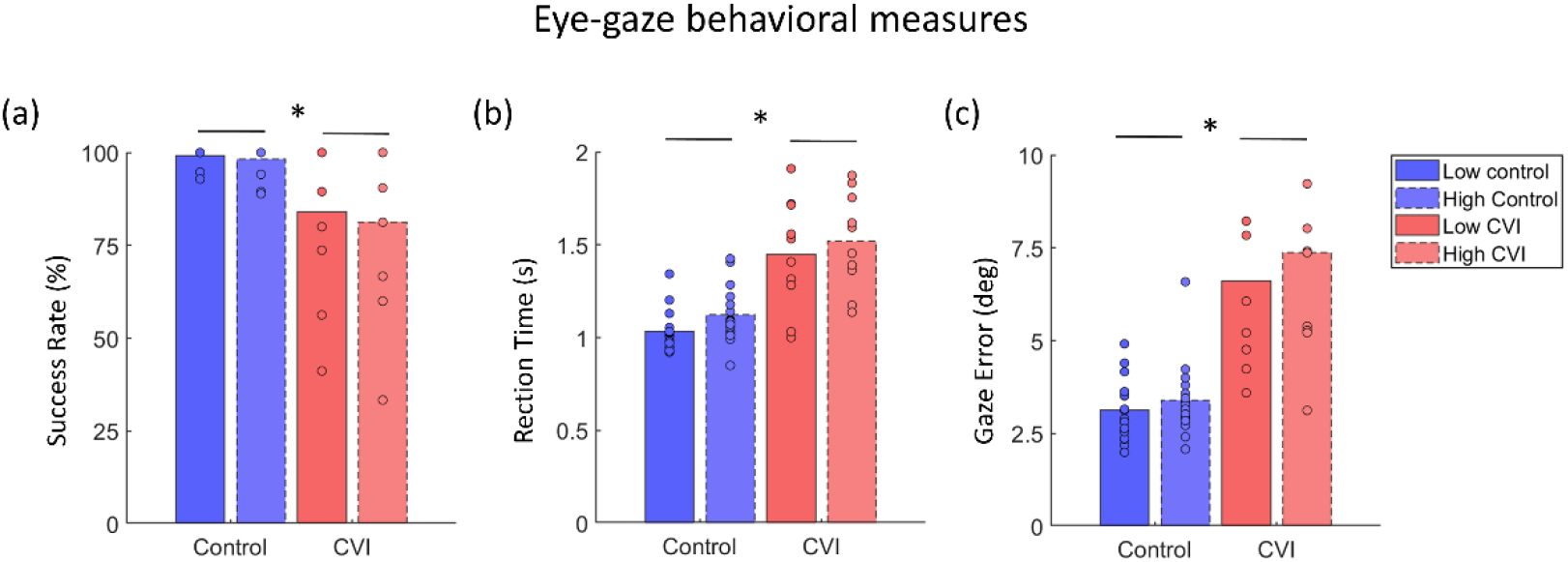
Behavioral Outcomes. Group bar plots for the outcomes of visual search performance (a) Success Rate, (b) Reaction Time, and (c) Gaze Error and with respect to task difficulty. CVI participants showed an overall profile of impaired visual search compared to controls.

Mean reaction times were higher in the CVI group than controls. A mixed ANOVA showed that the CVI group had a significantly higher reaction time (low: 1447.39 ± 297.85 msec, high: 1519.07 ± 259.22 msec) compared to controls (low: 1035.38 ± 109.54 msec, high: 1122.30 ± 151.59 msec) [F(1,24) = 29.27, p < 0.001, ηp2 = 0.55]. There was a significant effect of difficulty level [F(1,24) = 6.69, p = 0.016, ηp2 = 0.218], but no interaction effect [F(1,24) = 0.062, p = 0.806, ηp2 = 0.003] on reaction time. This suggests that the effect of task difficulty on reaction time was consistent across groups (figure 2 B). Finally, mean gaze error (an index for visual search accuracy) was significantly higher in the CVI group. A mixed ANOVA showed that the CVI group had a significantly higher gaze error (low: 6.61 ± 2.62 arc degrees, high: 7.37 ± 2.76 arc degrees) than controls (low: 3.14 ± 0.84 arc degrees, high: 3.39 ± 1.02 arc degrees) [F(1,24) = 27.275, p < 0.001, ηp2 = 0.532]. There was a significant effect of difficulty level [F(1,24) = 11.985, p = 0.002, ηp2 = 0.333], but no interaction effect [F(1,24) = 2.931, p = 0.100, ηp2 = 0.109]. This indicates that the effect of task difficulty on gaze error was consistent across groups (figure 3 C, see also supplementary figure S3 for representative examples of heat map displays of visual search patterns).

**Figure 3.**
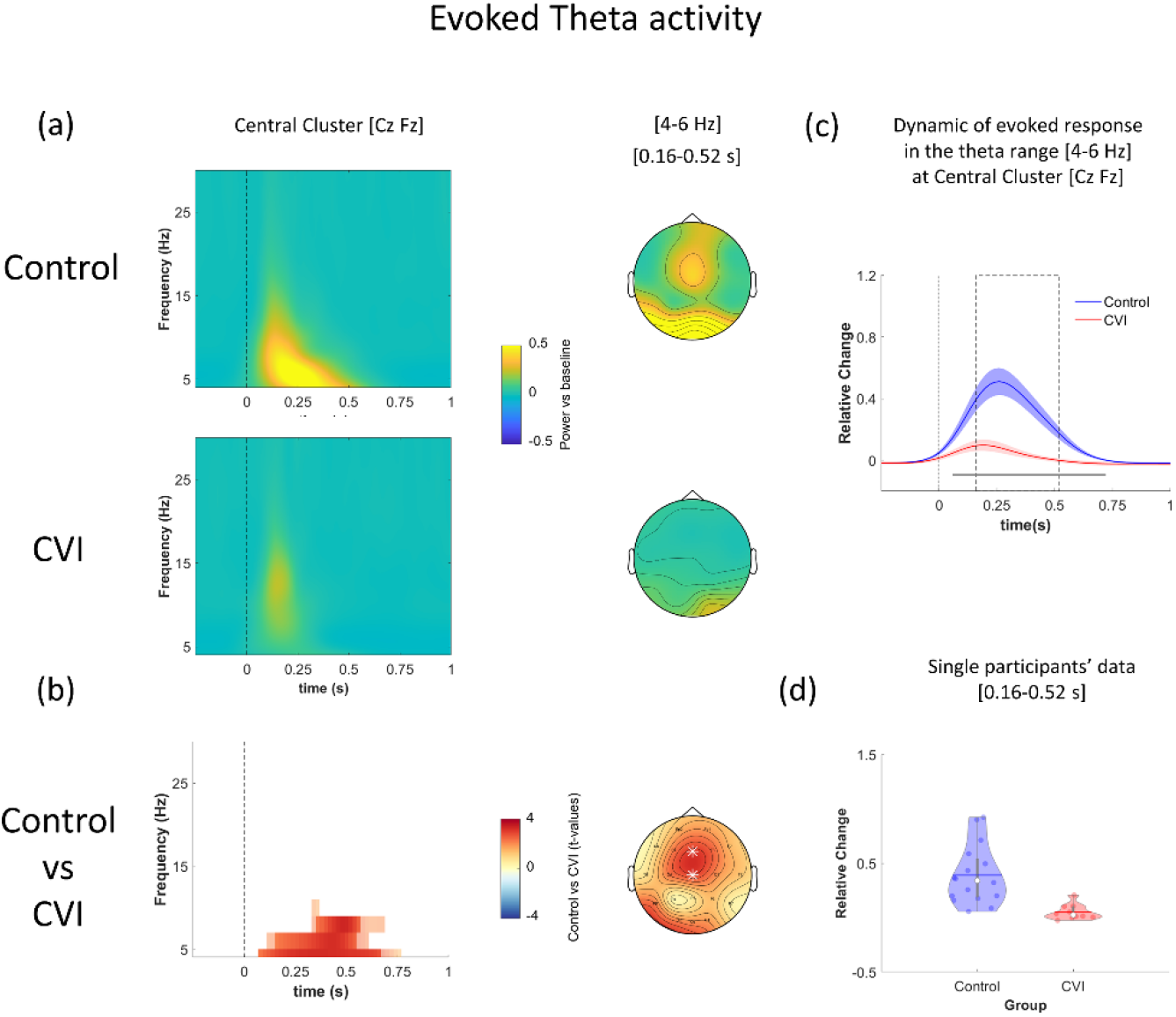
Evoked Theta Activity. (a) Oscillatory activity calculated as the average across low and high levels of task difficulty for controls (upper row) and CVI (bottom row) participants and plotted as a function of time [-0.25 - 1 s] and frequency [4-30 Hz]. The plots show the average between two representative central electrodes (Cz, Fz) and 0 sec indicates stimulus onset. Scalp topographies in the theta range [4-6 Hz] within a representative time window [0.16 - 0.52 sec] are shown (right panel). (b) Statistical results. Time-frequency plot highlighting significant differences between control and CVI participants identified by the cluster-based permutation test (p<0.025) and corresponding topography for theta range [4-6 Hz] at a representative time window [0.16 - 0.52 sec]. Electrodes belonging to the significant cluster are highlighted with white asterisks. (c) Time-course of the mean power in the theta range [4-6 Hz] for control and CVI subjects (data are averaged across two representative central electrodes Cz, Fz). Shaded areas represent SEM and the continuous horizontal grey line indicates the significant difference between CVI and control groups (from 0.08 to 0.72 s; FDR corrected). The dashed grey boxes represent the time window [0.16 - 0.52 sec] comprising the theta peak in which theta power [4-6 Hz] was extracted for each subject (across channels Cz, Fz) and shown in the corresponding violin plots (d) with each dot representing individual data.

Examining putative associations between visual acuity in CVI participants (based on the LogMAR value of the better-seeing eye) and all visual search outcomes did not reveal any significant statistical correlations (all p values > 0.24). Moreover, age did not emerge to be correlated with any behavioral measures of interest (all p values > 0.53).

Finally, we assessed whether behavioral performance was associated with structural morphometry data (MRI). We observed a significant negative correlation between gaze error and the volume of the pulvinar nucleus (r = −0.7167, p = 0.0298). However, this did not survive FDR correlation for multiple comparisons (p = 0.6258). There were no significant correlations between all the other behavioral outcomes and volume of the pulvinar, LGN, and pericalcarine cortex, respectively (all p values > 0.4).

### Neural Oscillations

#### Evoked power

Within the low frequency range [4-30 Hz], a cluster-based permutation test performed on the neural response change between the two levels of task difficulty and contrasting the control and CVI groups revealed an absence of a significant interaction (all p values > 0.03). Thus, we collapsed the low and high levels of task difficulty, and a cluster-based permutation revealed a significant difference between the two groups (p < 0.004). In controls, visual stimulus presentation elicited strong theta activity [4-6 Hz] between 150 and 500 msec, starting at occipital regions and extending to central areas. This pattern was nearly absent in the CVI group (see figure 3 A and B). No significant differences emerged at the high frequency range [30-55 Hz] (all p values > 0.1; see supplementary figure S4).

#### Induced power

Within the low frequency range [4-30 Hz], a cluster-based permutation test performed on the neural response change between task difficulty conditions and contrasting the control and CVI groups revealed a significant interaction between condition and group (p < 0.005). This interaction effect emerged within the alpha frequency band [8-14 Hz] at the occipital pole between 250 and 500 msec (see figure 4 A and B). We explored this further by performing a cluster-based permutation analysis between the low and high task conditions within each group. In the control group, we found a significant modulation of alpha (and also extending into beta) band activity associated with task difficulty (p < 0.001). In contrast, no difference emerged in CVI participants (all p values > 0.04). Moreover, when we investigated the difference between groups within each level of task difficulty, a cluster-based permutation analysis revealed a significant difference between the two groups for both low and high conditions (low: p < 0.001 and high: p < 0.005). In particular, the CVI group showed a reduction in alpha [10-16 Hz] desynchronization (> 500 ms) within occipital channels in the low condition (this difference also extended into beta activity [12-22 Hz] at the high level of task difficulty; see supplementary figure S5).

**Figure 4.**
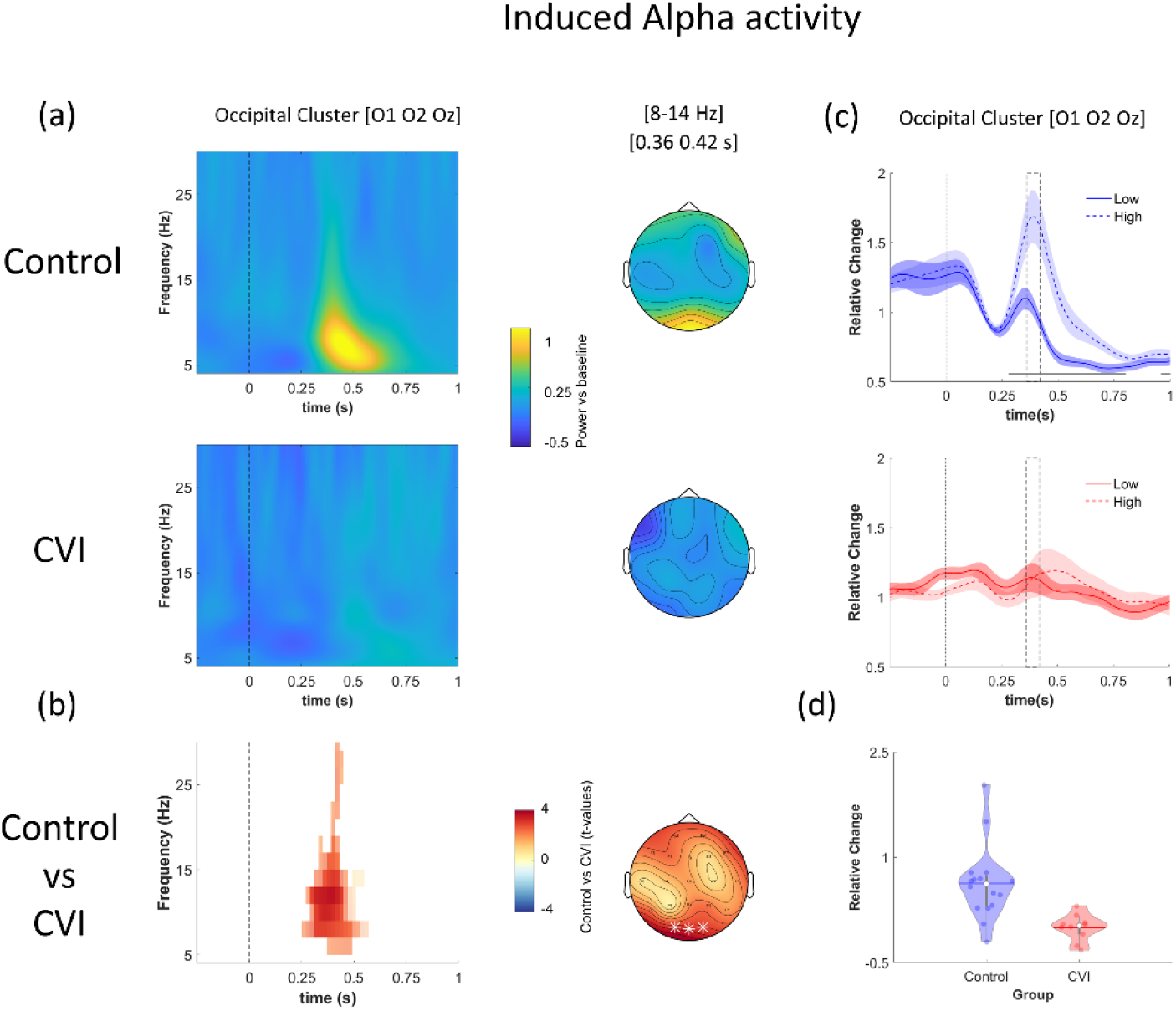
Induced Alpha Activity. (a) Oscillatory activity calculated as the difference between high and low levels of task difficulty is plotted for each group (controls: upper row and CVI: bottom row) as a function of time [- 0.25 - 1 sec] and frequency [4-30 Hz]. The plots show the average across three representative occipital electrodes (O1, O2, Oz) and 0 sec indicates stimulus onset. Scalp topographies in the alpha range [8-14 Hz] at a representative time window [0.36 - 0.42 sec] are shown (right panel). (b) Statistical results. Time-frequency plot highlighting significant differences between control and CVI groups identified by the cluster-based permutation test (p < 0.025) and the corresponding topography for the alpha range [8-14 Hz] at a representative time window [0.36 - 0.42 sec]; electrodes belonging to the significant cluster are highlighted with white asterisks. (c) Time-course of the mean power in the alpha range [8-14 Hz] separately for the low and high levels of task difficulty are shown for the control and CVI groups (upper and bottom rows respectively. Data are averaged across three representative central electrodes: O1, O2, Oz); shaded areas represent SEM; the continuous horizontal grey line indicates the significant difference between low and high levels of task difficulty in the control group (from 0.28 to 0.8 sec; p < 0.05, FDR corrected). The dashed grey boxes represent the time window [0.36 - 0.44 sec] comprising the alpha peak, in which the power in the alpha range [8-14 Hz] was extracted for each subject (across channels O1, O2, Oz) and shown in the corresponding violin plots (d), each dot represents individual data.

Within the high frequency range [30-55 Hz], a cluster-based permutation test performed on the neural response change between the two levels of task difficulty and contrasting the control and CVI groups revealed an absence of a significant interaction (all p values > 0.04). Thus, we averaged the individual oscillatory responses across the low and high levels of task difficulty, and a cluster-based permutation analysis revealed a significant difference between the two groups (p < 0.01). The effect emerged between 300 and 700 msec in the occipital-parietal area. In the CVI group, a delay in induced gamma [30-45 Hz] activity appeared to emerge compared to controls (see figure 5 A and B).

**Figure 5.**
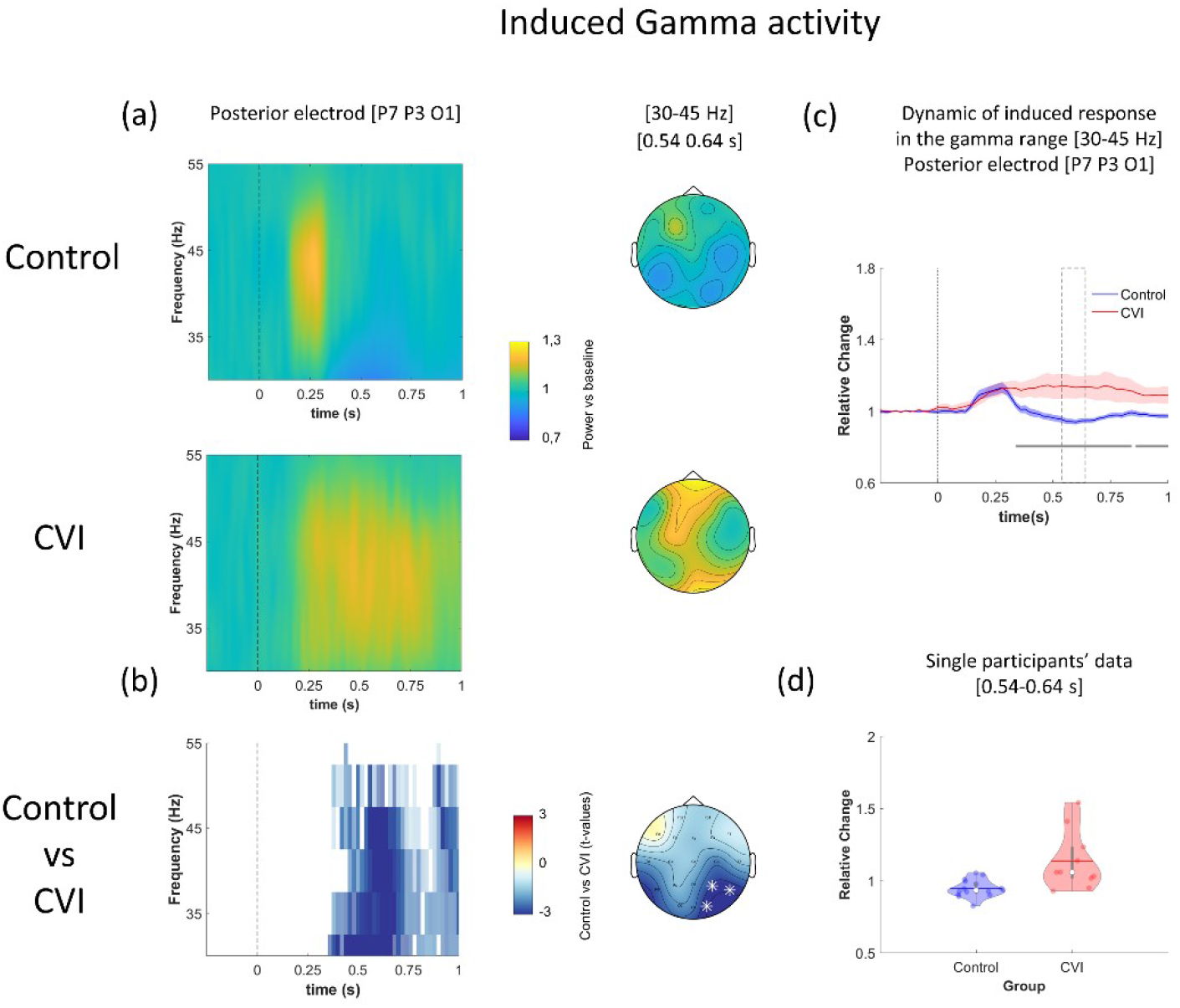
Induced Gamma Activity. (a) Oscillatory activity calculated as the average across low and high levels of task difficulty within each group (controls: upper row and CVI: bottom row) and plotted as a function of time [-0.25 - 1 sec] and frequency [30-55 Hz]. The plots show activity at representative posterior electrodes (P7; P3; O1) and 0 sec indicates stimulus onset. Scalp topographies in the gamma range [30-45 Hz] at a representative time window [0.54 - 0.64 sec] are shown in the right panel. (b) Statistical results. Time-frequency plot highlighting significant differences between control and CVI groups identified by the cluster-based permutation test (p < 0.025) and the corresponding topography for the gamma range [30-45 Hz] at a representative time window [0.54 - 0.64 sec]. Significant electrodes belonging to the significant cluster are highlighted with white asterisks. (c) Time-course at the group level of the mean power in the gamma range [30-45 Hz] for control and CVI groups (data averaged across representative posterior electrodes P7, P3, O1). Shaded areas represent SEM; continuous horizontal grey lines indicate significant differences between CVI and control groups (from 0.34 to 0.82 sec and from 0.86 to 1 sec; p < 0.05, FDR corrected). The dashed grey boxes represent the time window [0.54 - 0.64 sec] in which the power in the gamma range [30-45 Hz] was extracted for each subject (in the electrodes P7, P3, O1) and shown in the corresponding violin plots (d), each dot represents individual data.

Finally, to compare alterations in feedforward and feedback processing in CVI, we explored the relative strength of theta and alpha effects respectively. We found that the effect size was very strong and comparable for both theta (Cohen’s d = 1.55; CI: 0.66, 2.44) and alpha (Cohen’s d = 1.45; CI: 0.57 2.33).

### Event Related Potential (ERP) Analysis

No significant differences emerged in the ERPs (all p values > 0.034). However, visual inspection revealed a smaller P300 amplitude in the CVI compared to control group (see supplementary figure S6).

### Associations Between EEG Activity and Factors of Interest in CVI Participants

#### Relationship between EEG activity and volume of thalamic nuclei and primary visual cortex

There were no statistically significant correlations between the volumes of the LGN, pulvinar, pericalcarine cortex and average theta, alpha, or gamma activity nor P300 amplitude, respectively (all p values > 0.4).

#### Relationship between EEG activity and age, visual acuity, and global lesion classification

We did not observe any significant correlations between EEG activity (average theta, alpha, or gamma activity nor P300 amplitude) and age (all p values > 0.1), LogMAR visual acuity (all p values > 0.1), and extent of neurological damage (as defined by global lesion scores ^39^; all p values > 0.1).

## Discussion

We analyzed electrophysiological activity associated with higher-order visual perceptual impairments in individuals with CVI. At the behavioral level, CVI participants showed impaired visual search performance compared to controls as indexed by decreased success rate as well as increased reaction time and gaze error. This suggests that, in general, CVI participants were less likely, and took longer, in finding the target and that their search patterns were less accurate in finding the target compared to controls. Importantly, behavioral performance in CVI did not correlate with visual acuity, suggesting that impaired visual search could not simply be explained by reduced stimulus visibility, and highlights the brain-based nature of higher-order visual perceptual deficits associated with this condition. In general, these observations appear in line with the clinical profile of CVI with respect to visuospatial and visual attention processing deficits reported in this population ^4,8^.

The analysis of both evoked and induced oscillatory activity within a broad frequency range [4-55 Hz] revealed widespread alterations associated with impaired visual search in CVI participants compared to controls. Specifically, we found markedly reduced evoked theta [4-6 Hz] activity in the CVI group regardless of the level of task difficulty. Analysis of induced oscillatory activity revealed that alpha signal was also markedly reduced in CVI compared to control participants. Furthermore, while induced alpha activity in controls was enhanced at higher task difficulty, this modulation was not evident in the CVI group. Moreover, the CVI group showed a delayed, yet greater sustained induced gamma response compared to controls. These alterations in both evoked and induced components of neural oscillatory activity suggest that impaired visual search in CVI is associated with anomalies in both feedforward and feedback signaling.

### Time and Phase-Locked Neural Oscillations Associated with Feedforward Processing

The analysis of evoked oscillatory activity revealed a significant difference between CVI and control groups with respect to theta activity. In controls, a robust post-stimulus response in evoked theta [4-6 Hz] was found within the first 500 msec of visual stimulus presentation. However, in CVI, this theta activity was very much reduced, and independent of the level of task difficulty. Theta activity has been linked with the active sampling of input from the environment, with information processed at any given moment packed in each theta cycle ^59^. Relevant to this study, successful visual search was found to be positively associated with post-stimulus theta amplitude in normally sighted individuals ^60^. Interestingly, a subset of our study participants with CVI had visual acuities within normal/near normal levels yet, impaired theta activity was still evident. This suggests that a reduction in evoked theta activity could be linked to a specific deficit in the organization of the neural response time phase-locked to visual events. In other words, even in the case of CVI participants having normal visual acuity, feedforward visual processing appears to be impaired in the setting of early neurological injury.

### Non Time and Non Phase-locked Neural Oscillations Associated with Feedback Processing

Analyzing induced oscillatory activity, we found evidence of two major alterations in the CVI group associated with alpha [8-14 Hz] and gamma [30-45 Hz] activity. In controls, occipital induced alpha activity increased with higher visual task difficulty. However, this signal modulation was absent in CVI participants. Given the well-established link between alpha activity and visual attention ^38,61-63^, this finding would be consistent with the view that neural mechanisms implicated with selecting visually attended targets and suppressing task-irrelevant stimuli are impaired in CVI ^64,65^. A recent study by Gutteling and colleagues reported that oscillations in the alpha band may be especially involved in distractor suppression and play a key role in closing the perceptual gate to distractor input ^66^. In the context of the results presented here, it is possible that CVI participants may have specific deficits with the inhibition of surrounding distracting information.

In addition, CVI participants showed a delayed yet, sustained induced gamma activity within posterior regions of the scalp and independent of the level of task difficulty. Gamma activity has been consistently associated with ocular movements such as micro-saccades ^67^. Thus, it is possible that this aberrant pattern in gamma oscillatory activity in CVI could also be related to altered visual search behavior. Consistent with this view, our visual search data suggest that gaze error (an index of gaze accuracy with respect to locating and accurately fixating the target) was significantly higher in CVI compared to control participants. The continuous need to recalibrate gaze while searching, locating, and fixating the target appears consistent with the pattern of sustained gamma band activity observed in CVI.

### Neural Markers of CVI and their Potential Clinical Significance

To our knowledge, studies investigating electrophysiological activity (beyond VEP recordings) in relation to higher order visual processing in CVI remain limited. A recent study by verMass and colleagues (2021) investigated children with CVI associated with cerebral palsy (CP) and used magnetoencephalography (MEG) to uncover oscillatory activity associated with visuospatial processing abilities ^68^. Consistent with the findings reported here, the authors found that participants with CP had weaker theta (as well as gamma) occipital activity related to impaired performance on their visuospatial processing and attention task. Consistent with our findings, the authors proposed that these changes in oscillatory activity may be related to poor bottom-up processing of incoming visual information that in turn impacts higher-order visual processing and decision making ^68^. In our study, not all of our participants with CVI were diagnosed with CP nor had underlying neurological damage associated with PVL. Thus, our results further extend these observations to a broader profile of CVI as well as in relation to impaired visual search performance across multiple etiologies associated with this condition.

Our analysis of oscillatory activity in CVI revealed marked alterations with respect to feedforward processing, as suggested by the nearly absent evoked responses in the theta range. Moreover, widespread signal alterations in CVI group were also evident with respect to induced oscillatory activity. Our findings appear to indicate that early onset neurological damage impacts the correct development of feedback processing as well. The identification of cerebral structures implicated with altered feedforward and feedback processing remain unclear but may be related to injury along thalamo-cortical visual pathways. In line with this view, we observed a significant negative correlation between gaze error and the volume of the pulvinar nucleus. This appears consistent given the important role this nucleus plays in the control of saccadic eye movements and the modulation of visual attention ^69,70^ (see also ^71^ for discussion of the pulvinar in distractor processing and visual search). Thus, the relative combination of aberrant feedforward and feedback processing may represent a distinctive marker of CVI in relation to impaired visuospatial processing with respect to visual search abilities.

Normal development of feedforward connectivity is crucial for stimulus driven sensory processing and seems to precede the elaboration of feedback connectivity, which in turn is important for top-down control of sensory processing and learning ^72,73^. It has been suggested that early stimulus-driven learning plays a crucial role in elaborating feedback connectivity ^74,75^. The present results suggest that cortico-cortical feedback connectivity not only needs external input, but also requires intact central processing to fully develop and operate.

### Study Limitations and Future Considerations

Electrophysiological data in this study were recorded using a simple 20-channel EEG system to facilitate acquisition in the clinical setting and minimize participant discomfort (note that eye movements were also recorded with eye tracking). Given the reduced number of electrodes employed, we could not perform source modeling to localize neural sources of the electrophysiological activity. Future studies using our visual search paradigm combined with high-density EEG or MEG are therefore needed to further characterize the nature of associated neural activity.

The relatively small sample size of our CVI group also represents an important limitation and is related to the challenges of subject recruitment for these types of studies. Specifically, due to the technical demands of this study, we recruited CVI participants within a limited range of visual functioning (i.e., visual acuity, visual field and contrast sensitivities, as well as ocular motor functions needed to maintain fixation) so as to minimize the effect of potential confounding factors that could influence visual search performance. Thus, this relatively narrow profile may limit the generalizability of our results to the overall CVI population. Future studies using other behavioral assessment designs that can reveal a more robust profile relating performance and task difficulty in combination with EEG recordings could help to better characterize associations between impaired behavioral responses and oscillatory activity. Finally, we observed deficits of both feedforward (theta) and feedback (alpha) processing in the CVI group. However, data showed that their functional role was not the same. Only neural oscillations in the alpha range increased with task difficulty in controls. Importantly, this modulation was absent in CVI participants, indicating a specific role of this oscillatory activity in visual search behavior and distraction suppression. Future studies may consider investigating relative contributions of feedforward and feedback activity in other visual contexts to understand their value as a potential biomarker and potentially help inform the design of specific training strategies.

## Conclusions

In conclusion results from this study suggest that participants with CVI have impaired visual search performance, consistent with previous accounts related to impaired visuospatial and visual attention processing. Impaired visual search performance was associated with marked alterations in neural oscillatory activity over a broad frequency range. This was consistent with widespread deficits within the visual system and implicating both local and distributed levels of neural processing. Finally, alterations of both evoked and induced components of oscillations suggest that both feedforward and feedback processing may be compromised in CVI.

## Data Availability

All data produced in the present study are available upon reasonable request to the authors

## Abbreviations

CVI: Cerebral Visual Impairment
ERP: event-related potential
LGN: lateral geniculate nucleus
VEP: visual evoked potential

## Acknowledgements

The authors would like to thank the subjects and their families for their participation in this study.

## Supplementary material

### Diagnosis of CVI

Diagnosis was based on a directed and objective assessment of visual functions including visual acuity, contrast, visual field perimetry, color, and ocular motor functions. A thorough refractive and ocular examination was carried out, as well as an extensive and integrated review of medical history (including developmental, birth, and gestational), neuroimaging, and electrophysiology records ^8,76,77, see also {Boonstra, 2022 #178^}. Details regarding visual behaviors were collected using standardized questionnaires and inventories ^78,79^.

### Development of Visual Environment and Eye Calibration

The visual environment was developed using the Unity 3D game engine (version 5.6; Unity Technologies) on an Alienware Aurora R6 desktop computer (Intel i5 processor, NVidia GTX 1060 graphics card, 32 GB of RAM; Alienware Corporation). In house 3D object models were created using Blender modeling software (Blender Foundation). The visual search task was run using Presentation software (https://www.neurobs.com) to control simultaneous EEG event markers and signal recording.

Eye tracking data (corresponding to X and Y coordinate positions of gaze on the screen) were captured using a Tobii 4C Eye Tracker system (90 Hz sampling frequency, Tobii Technology AB, Stockholm, Sweden). Eye tracking calibration was performed on each participant prior to data collection (Tobii Eye Tracking Software, v 2.9 calibration protocol). This included a 7-point calibration task (screen positions: top-left, top-center, top-right, bottom-left, bottom-center, bottom-right, and center-center) followed by a 9-point post calibration verification (i.e., the same 7 calibration points as well as center-left and center-right positions). Accuracy criterion was determined by gaze fixation falling within a 2.25 arc deg radius around each of the 9 points and was further confirmed by visual inspection prior to data collection.

**Supplementary Figure S1.**
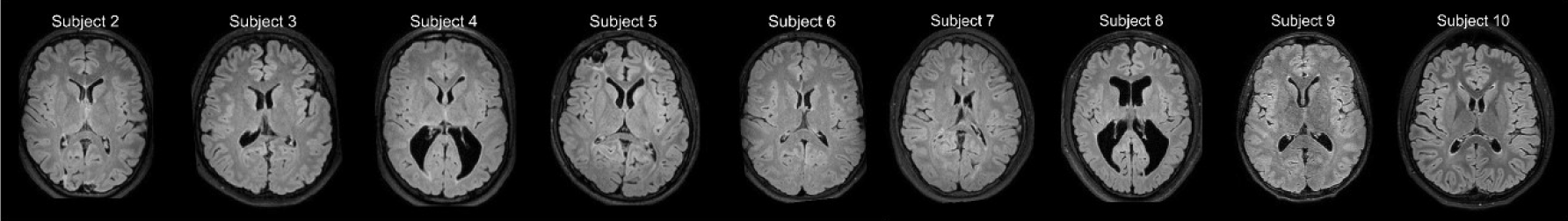
MRI. Axial fluid-attenuated inversion recovery (FLAIR) images from CVI study participants (scanning could not be performed on participant 1 due to contraindications to MRI). Representative images illustrate examples of the underlying neurological injury in CVI such as periventricular leukomalacia (PVL), white matter injury, and cortical atrophy.

**Supplementary Figure S2.**
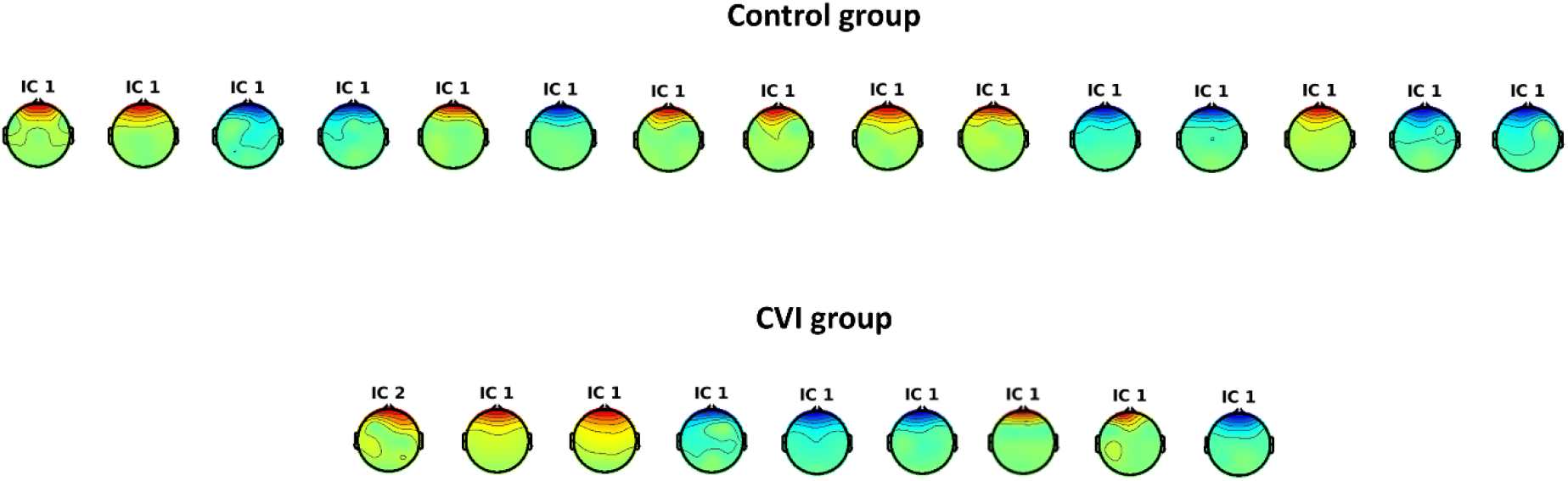
Removed Blink Components. Topography of each blink component removed from each participant (first; IC 1 or second; IC 2). Only 2 participants (1 control and 1 CVI) did not show typical blink artifacts.

**Supplementary Figure S3.**
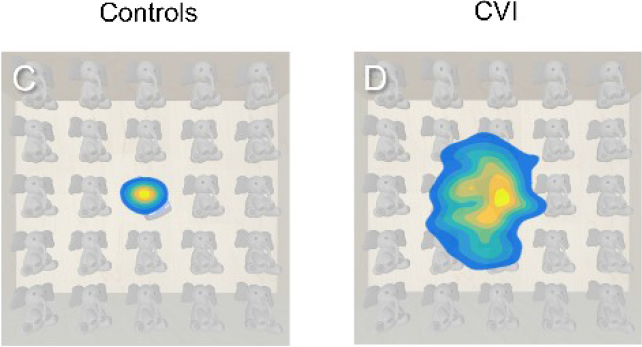
Heat Map Displays of Visual Search Patterns. Captured eye tracking data were aggregated and centered over time to generate heat maps representing the spatial extent of visual search patterns in each participant. The color scheme represents differing levels of gaze data density across spatial regions of the screen space (yellow indicates more time looking in an area and blue indicates less time). A) Data from a control participant shows a tight clustering of eye movements around the target while in a CVI participant (B), there is an overall larger distribution of gaze points around the target.

**Supplementary Figure S4.**
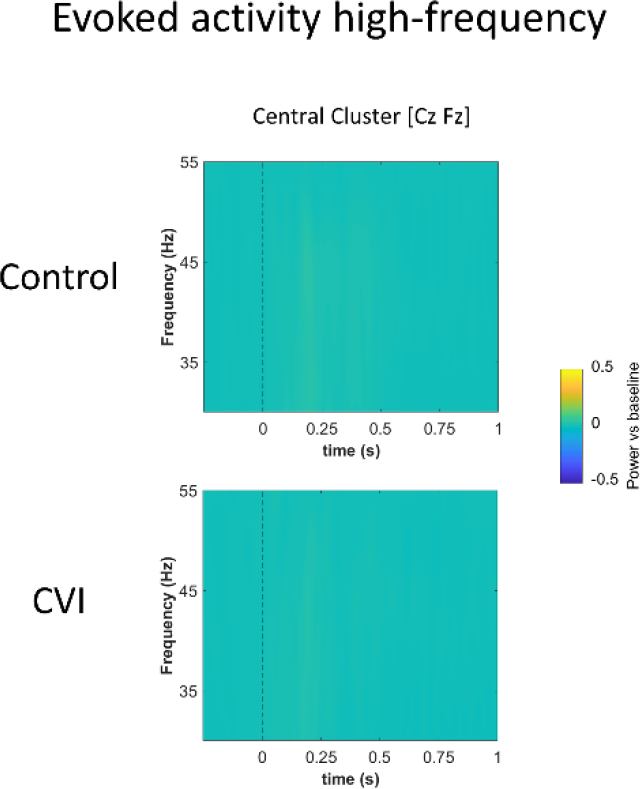
Evoked Activity in the High Frequency Range. Oscillatory activities calculated as the average across low and high levels of task difficulty within each group (controls: upper row and CVI: bottom row) as a function of time [-0.25 - 1 sec] and frequency [30-55 Hz]. The plots show the average between two central electrodes (Cz, Fz) and 0 sec indicates stimulus onset.

**Supplementary Figure S5.**
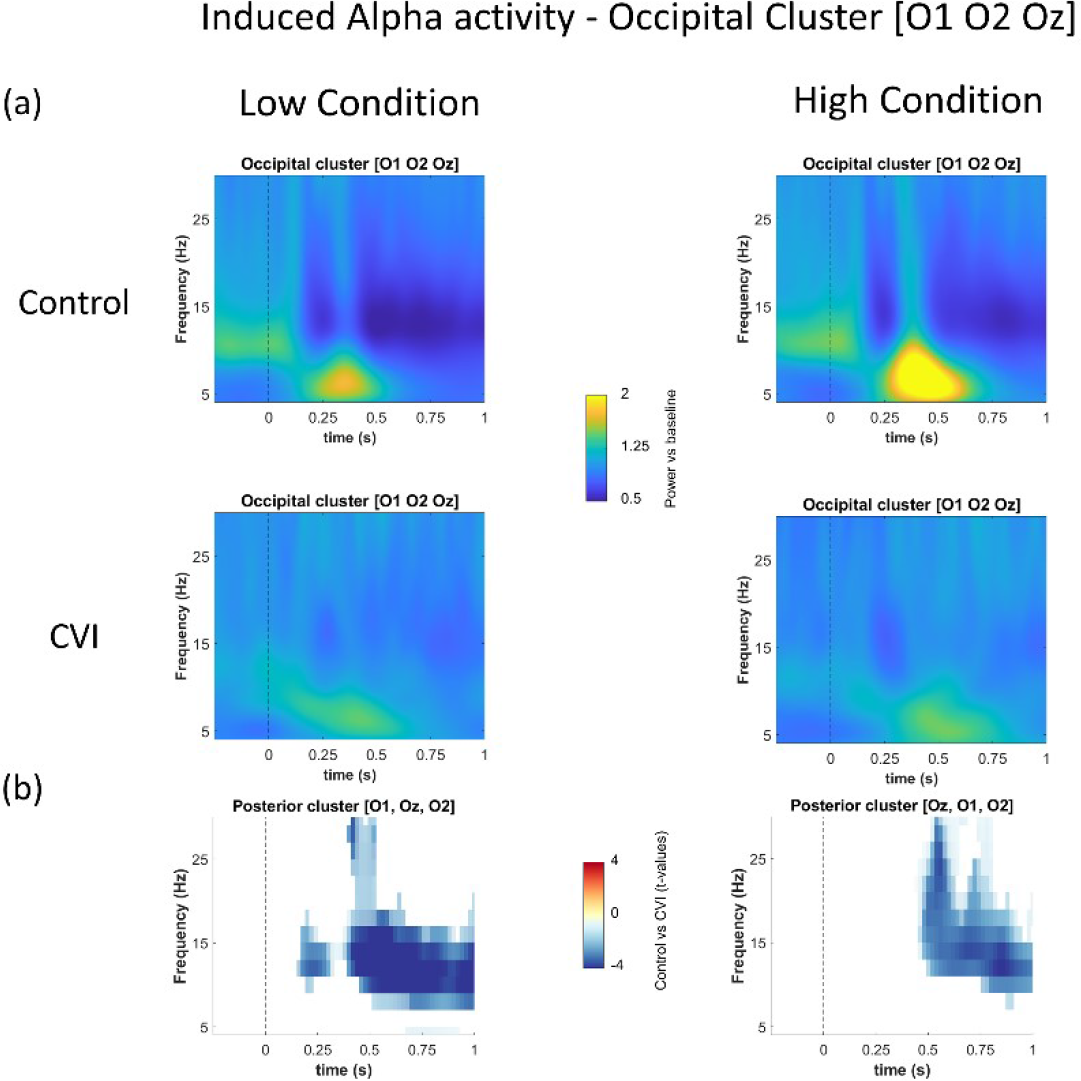
Induced Alpha Activity Associated with Low and High Task Difficulty. (a) Activity is plotted within each group (controls: upper row and CVI: bottom row) and within each condition (low task difficulty: left column and high task difficulty: right column) as a function of time [-0.25 - 1 sec] and frequency [4-30 Hz]. The plots show the average across three occipital electrodes (O1, O2, Oz) and 0 sec indicates stimulus onset. (b) Statistical results in the low (left column) and high (right column) conditions. Time-frequency plot highlighting significant differences between the control and CVI groups identified by the cluster-based permutation test (p<0.05, two-tailed).

**Supplementary Figure S6.**
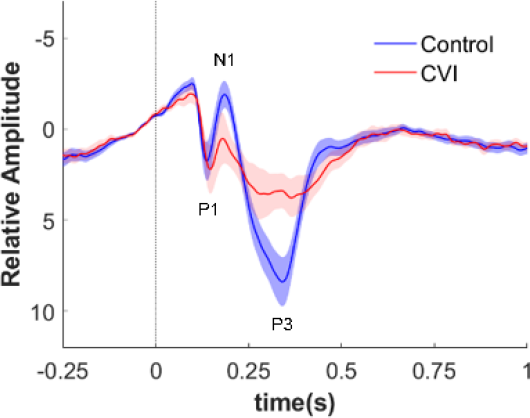
Event Related Potential (ERP). One-tailed positive cluster-based permutation performed on posterior electrodes (P8, P4, Pz, P3, P7, O1, Oz, and O2) within [0-0.5 sec] revealed a significant difference between 300-400 msec (p<0.045) consistent with decreased P300 amplitude in CVI subjects compared to controls.

